# Post-Acute COVID Syndrome, the Aftermath of Mild to Severe COVID-19 in Brazilian Patients

**DOI:** 10.1101/2021.06.07.21258520

**Authors:** Ana Paula Andrade Barreto, Lucimeire Cardoso Duarte, Thiago Cerqueira-Silva, Marcio Andrade Barreto Filho, Aquiles Camelier, Natalia Machado Tavares, Manoel Barral-Netto, Viviane Boaventura, Marcelo Chalhoub Coelho Lima, on behalf of the CPC study group

**Author notes:** Correspondence: Marcelo Chalhoub Coelho Lima, Address: Hospital Especializado Octávio Mangabeira, Praça Conselheiro João Alfredo, S/N, Pau Miúdo, Salvador- BA, 40320-350. These authors contributed equally to this work (Ana Paula Andrade Barreto, Lucimeire Cardoso Duarte, Viviane Boaventura, Marcelo Chalhoub Coelho Lima).

## Abstract

**Objective:** To describe persistent symptoms after acute COVID-19 in different spectrum of disease severity in a population from an upper/middle income country, and identify the main clinical features impacting the quality of life.

**Design:** Cross-sectional study.

**Setting:** Outpatient clinic from a public post-COVID-19 health center (CPC) at Bahia-Brazil, a state where 80% are black or mixed race.

**Participants:** Patients admitted between August 2020 and February 2021 with symptoms at least one month after the onset of COVID-19.

**Main outcome measures:** PACS and related disorders such as hospitalization one month or later after disease onset, biochemical dysregulation and reduced quality of life (EQ-5D-5L questionnaire).

**Results:** Among 683 individuals assisted at CPC in this period, 602 were recruited. Patients had average of 52 (±14.6) years, 355 (59%) were female, 528 (88%) black/brown. Individuals were classified as mild (39.9%), moderate (27.9%) or severe (32.2%) during acute illness if outpatient, hospitalized non-UCI or UCI, respectively. Most patients reported a polysymptomatic profile, in median eight (IQR=6-9) acute symptoms. The most frequent residual symptoms were dyspnea (66%), fatigue (62%) and chest pain (43%). Women were more affected regardless disease severity at acute stage: presented more residual symptoms [4 (2-6) vs 3 (2-4)] and a higher impact in quality of life. Altered HbA1c [(184/275 (66.9%)], high CRP levels [195/484 (40.3%)] and anemia [143/545 (26.2%)] were the most common abnormalities in laboratory exams. 76 patients presented HbA1c above 6.4% although only 42 referred previous diagnosis of diabetes mellitus. After one month of disease onset, 30 patients required hospitalization, including seven cases with mild acute illness. Hospital admission after acute disease was required on 30 patients, seven (23%) were mild. Quality of life had been affected for 357/404 (88.4%) patients according to EuroQoL (EQ-5D-5L), mainly the domains of anxiety/depression [severe or extreme anxiety for 79/401 (19.7%)] and pain/discomfort [severe or extreme pain for 71/403 (17.6%)]. The median EuroQoL Global Score was 70 [IQR 50-80]. PACS symptoms such as dyspnea, chest pain, and fatigue, was associated with decreased quality of life.

**Conclusions:** PACS, such as dyspnea, chest pain and fatigue, occurred after variable degree of disease severity. Among this majority black/mixed-race patients, woman seemed to be more affected. Other consequences included post-acute hospitalization, and abnormal glucose metabolism and reduced quality of life.

**Summary Box:** *Section 1: What is already known on this topic:*

✓ Post-Acute COVID Syndrome (PACS) comprises a set of persistent or new-onset symptoms after illness onset.
✓ As far as we know, there are no studies describing PACS in a population principally black and mixed-race. Additionally, few studies have addressed PACS among outpatients.

*Section 2: What this study adds:*

✓ Similar PACS were reported after mild, moderate and severe illness. Dyspnea, fatigue and chest pain were the most prevalent symptoms in this population presenting majority of black/mixed-race patients.
✓ Women presented more residual symptoms, a higher frequency of myalgia and worse score for mobility, usual activities, anxiety/depression, and pain.
✓ Hospitalization may occur one month or later after mild or moderate/severe acute infection due to respiratory and vascular disorders. Abnormal glucose metabolism was detected in the absence of previous diagnosis of diabetes mellitus.

## INTRODUCTION

One year after COVID-19 was declared a pandemic, more than 141 million people have been infected, many of whom persist with symptoms[1,2]. This clinical condition, denominated Post-Acute COVID Syndrome (PACS), involves early and late-onset symptoms, including respiratory, musculoskeletal, and sensorineural complaints detected no less than four weeks after the onset of disease [2].

PACS has been mainly reported after hospitalization[3,4], and may be linked to post-intensive care syndrome and post-traumatic stress disorders. Few studies have addressed the persistence of symptoms after mild disease[5–7]; six months after the onset of COVID-19, approximately one-third of these patients remain symptomatic. Moreover, most reports on the effect of PACS on patients’ quality of life focus on European and North American regions [7].

COVID-19 disproportionately affects the most vulnerable communities[8,9]. While Brazil is an upper-middle income country, and one of the countries most severely affected by COVID-19, with almost 14 million confirmed cases as of April 2021[1], the long-term consequences in this population have not been investigated.

Here, we sought to characterize patients from Brazil with PACS, focusing on the full spectrum of disease presentation during the acute phase. We analyzed persistent symptoms according to gender and disease severity and identified the main clinical features impacting these patients’ quality of life.

## METHODS

### Study design, population and ethical aspects

We report a cross-sectional study that provides the baseline characteristics of a cohort of PACS cases seen at Centro Pós-Covid-19 (CPC), a public health outpatient clinic that operates at the Octávio Mangabeira Specialized Hospital (HEOM), located in Salvador-Bahia, Brazil. CPC provides outpatient services to residents of the state of Bahia (15.13 million inhabitants).

Patients were recruited by invitation upon discharge from local COVID-19 reference hospitals, or came to CPC on an outpatient basis (advertisements were circulated via conventional and social media). The inclusion criteria were: 1) Recruitment at least one month after disease onset; 2) positive SARS-CoV-2 reverse transcriptase–polymerase chain reaction (RT-PCR) test, or IgM/IgG serological test positivity, or a computed tomographic (CT) lung scan compatible with viral pneumonia in association with clinical features. Exclusion criteria consisted of: age <18 years, cognitive disorders, pregnancy or a lack of residual symptoms.

The present study was approved by the institutional review boards of the Bahia State University (UNEB; protocol no. 38281720.2.0000.0057) and the Santo Antonio Hospital (OSID; protocol no. 33366030.5.0000.0047).

### Clinical and laboratorial evaluations

Upon recruitment at CPC, patients were routinely evaluated by a multidisciplinary team (consisting of a nurse, physician, physiotherapist, nutritionist, psychologist and social worker). Patients were classified according to disease severity during the acute phase of infection (within the first 30 days following disease onset): mild (not requiring hospitalization), moderate (non-ICU hospitalization) or severe (ICU admission). Data were collected and managed using Research Electronic Data Capture (REDCap) software, with hosting provided by the Gonçalo Moniz Institute (IGM-FIOCRUZ), located in Bahia, Brazil[10,11]. Sociodemographic characteristics were recorded, in addition to anthropometric parameters, oxygen saturation, comorbidities, social habits, clinical features, current/past treatments and hospitalization post-acute phase (i.e., more than one month of disease onset). Patients answered a structured survey to provide detailed information on manifestations during the acute phase of infection, as well as the persistence of/ onset of new symptoms post-acute phase. Additionally, quality of life was assessed using the Portuguese version of the EuroQol (EQ-5D-5L) instrument, which evaluates mobility, self-care, usual activities, pain, anxiety/depression and general health status[12]. Blood samples were collected to evaluate the following laboratory parameters: complete blood count, glycemia, hemoglobin A1C (HbA1c), urea, creatinine, AST, ALT, bilirubin, C-reactive protein, sodium, potassium and creatine kinase. Due to technical issues, HbA1c was only available in 46% of the included patients.

### Statistical analysis

All data were analyzed using the R software package [13]. Continuous variables with normal distributions were described as means and standard deviation, whereas variables with non-normal distribution were described as medians and interquartile range. Categorical variables were expressed as frequencies and percentages. Data distribution was assessed using the Shapiro-Wilk test and by histogram analysis. Comparisons between quantitative variables in two independent groups were made using the Student’s T or Mann-Whitney tests; for three or more groups, we used ANOVA or Kruskal-Wallis tests. The Chi-square test was used to assess differences between categorical variables. Ordinal logistic regression analysis was performed for each domain of the EuroQol, considering age, sex, BMI, hospitalization during the acute phase, and the three most prevalent persistent symptoms. Linear regression was carried out using global EuroQol scores. The Brant test was used to test for proportionality [14].

## RESULTS

From August 2020 to February 2021, 683 patients with confirmed or suspected COVID-19 were seen at CPC. Patients were excluded when no residual symptoms were presented/reported at the initial appointment (n=50), if they refused to participate (n=23), presented no confirmatory diagnosis of COVID-19 (n=5) or were considered ineligible due to pregnancy or severe psychiatric illness (n=3) (Figure 1). The remaining 602 individuals (mean age: 52 years, 59% female) were classified as having mild (39.9%), moderate (27.9%) or severe (32.2%) disease during the acute phase of infection. The patients seen at CPC mostly self-reported skin color as mixed-race or black (52.3% and 35.4%, respectively). Most patients reported at least one comorbidity (70.4%), mainly hypertension (43.1%) followed by obesity (40.2%). Among the moderate/severe cases (n=362, 60.1%), almost half were women (48.1%) who presented a significantly higher BMI than the men in this same classification (31.7±7.7 vs 28.7±4.9, p<0.001). Of the 479 patients who responded to a question on weight loss during the acute phase of disease, 276 (57.6%) reported weight loss and 182 (66.4%) of these were not obese at the time of their initial appointment at CPC. Most patients reported polysymptomatic acute infection, with a median of eight symptoms (IQR=6-9), including respiratory complaints (82% dyspnea, 77% cough), fatigue (81%), myalgia (70%), headache (68%), chest pain (62%), fever (62%) and olfactory and gustatory dysfunction (60%) (Table 1 and Supplementary Table 1).

**Table 1.** Demographics, clinical characteristics and acute phase symptoms of PACS patients.

|  | **Overall (N=602)** |
| --- | --- |
| **Age, years** | 51.8 (±14.6) |
| **Female** | 355 (59.0) |
| **Ethnicity** |  |
| Mixed-race | 315/601 (52.4) |
| Black | 213/601 (35.4) |
| White or other | 73/601 (12.2) |
| **BMI-kg/m<sup>2</sup>** | 29.6 (±6.3) |
| **Time of disease onset (months)** | 2.5 (1.6 - 4.2) |
| **Diagnostic\*\*** |  |
| **RT-PCR** | 460 (76.4) |
| **Serologic (IgM/IgG)** | 31 (5.1) |
| **Point of care - Serologic** | 92 (15.3) |
| **Point of care - Antigen** | 25 (4.2) |
| **Radiologic + Clinical** | 12 (2.0) |
| **Any comorbidity** | 424 (70.4) |
| **Hypertension** | 259/601 (43.1) |
| **Obesity** | 241/599 (40.2) |
| **BMI range- kg/m<sup>2</sup>\*** |  |
| **30-34** | 154 (26.0) |
| **35-39** | 52 (8.8) |
| **>40** | 34 (5.7) |
| **Diabetes Mellitus** | 141/601 (23.5) |
| **Asthma** | 65/599 (10.9) |
| **Cardiovascular disease** | 51/598 (8.5) |
| **Chronic Obstructive Pulmonary Disorder** | 23/594 (3.9) |
| **Other (Chronic Kidney Disease, Neoplasia)** | 20/599 (3.3) |
| **Cigarette smoking (current/former)** | 154/596 (26.2) |
| **Alcoholism** | 8/594 (1.3) |
| **Number of symptoms** | 8.0 (6.0 - 9.0) |
| **Fever** | 370/600 (61.7) |
| **Cough** | 461/601 (76.7) |
| **Myalgia** | 422/601 (70.2) |
| **Headache** | 411/601 (68.4) |
| **Diarrhea** | 238/601 (39.6) |
| **Dyspnea** | 492/601 (81.9) |
| **Vomiting/Nausea** | 199/601 (33.1) |
| **Olfactory dysfunction** | 360/600 (60.0) |
| **Gustatory dysfunction** | 362/601 (60.2) |
| **Fatigue/Muscle weakness** | 484/601 (80.5) |
| **Chest pain** | 371/601 (61.7) |
| **Glucocorticoid use** | 357/493 (72.4) |
| **Weight loss** | 276/479 (57.6) |

Data are n (%), n/N (%), mean ( ± SD) or median (IQR)
**\*n=599**
**\*\*Some patients were diagnosed through more than one test.**

**Figure 1.**
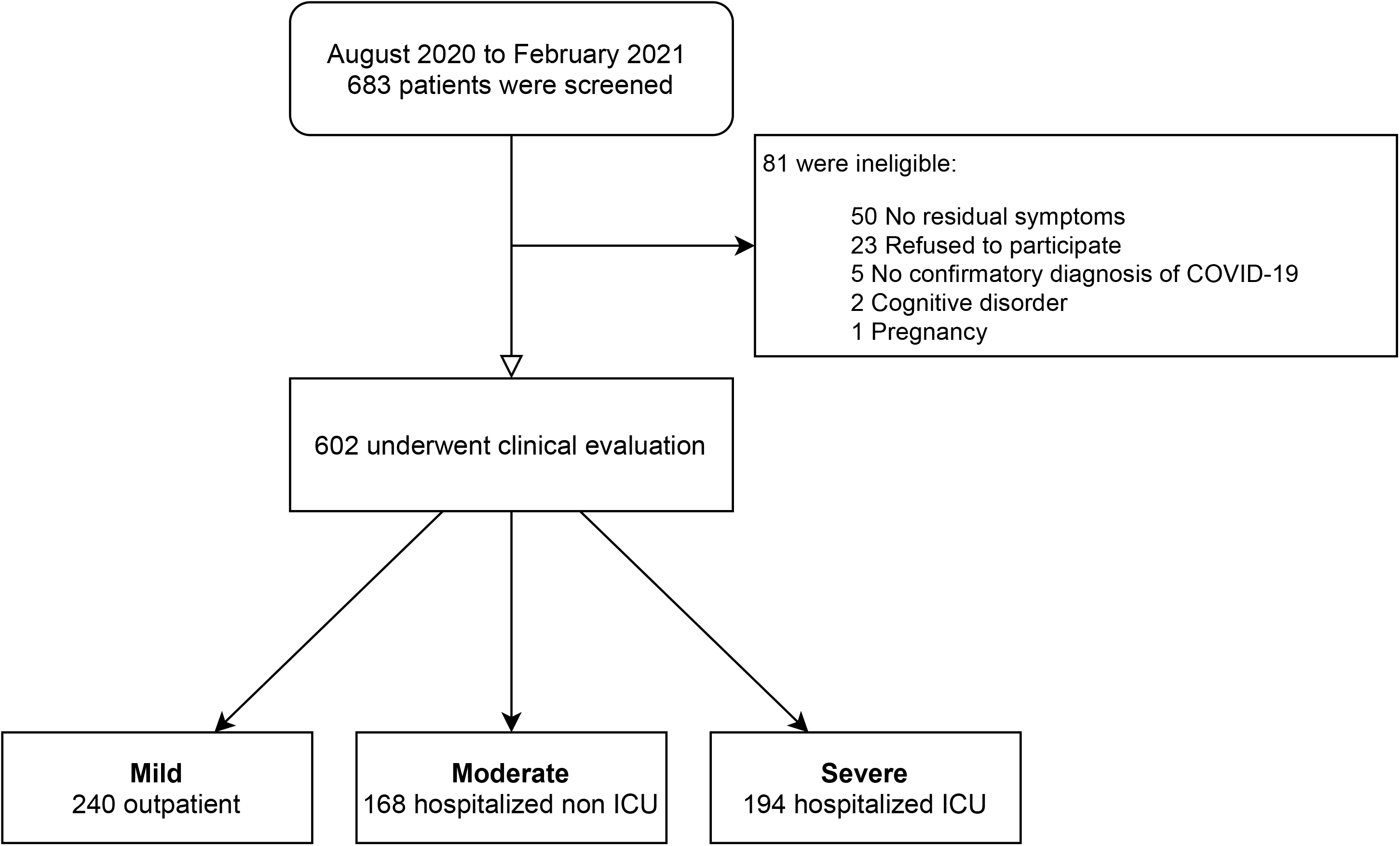
Flowchart of patients with PACS attended at CPC - Bahia - Brazil.

The patients were evaluated at a median of 2.6 [IQR=1.6-4.3] months after the onset of disease. A greater number of persistent symptoms were noted in women post-acute infection, as well as a higher frequency of myalgia across all disease severity groups (Figure 2A and Supplementary Table 1). Dyspnea was the most prevalent PACS-associated symptom, with more than 60% reporting it across all disease severity strata; 43% of these individuals had a modified Medical Research Council (mMRC) Dyspnea score ≥2. Fatigue (61.6%) and chest pain (43.4%) were also frequently reported (Supplementary Table 1). The persistent symptoms reported in the context of PACS usually initiated during the acute infection. Late-onset chest pain (13.6-25%), myalgia (14.3-21%) and fatigue (9-16.8%) were reported at variable rates in accordance with disease severity (Figure 2B).

**Figure 2.**
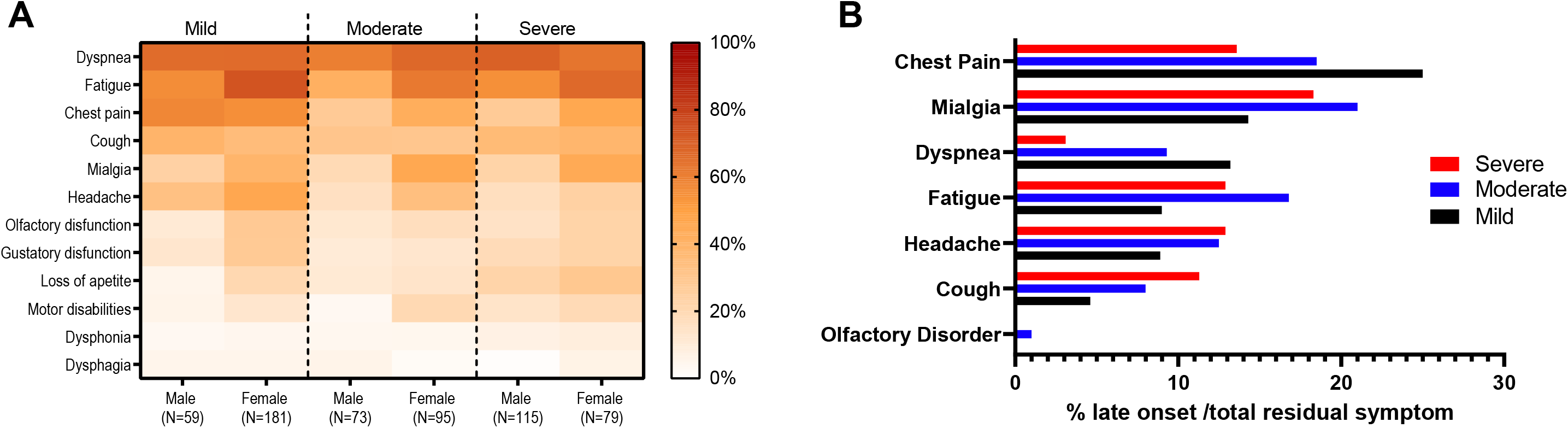
Frequency of residual symptoms among patients PACS patients. A- Heat map illustrating the frequency according to sex and disease severity. B- Percentage of late-onset symptoms according to disease severity.

Increased levels of CRP [195/484 (40.3%)] was the most common alteration on laboratory exams, followed by anemia in 143/545 (26.2%). HbA1c testing was available for 275 patients, with 184 (66.9%) presenting elevated values. HbA1c levels were above 6.4% in 76 patients (27.6%), and 34 of these individuals had no previous diagnosis of diabetes mellitus (DM). The remaining 108 (39.3%) patients presented HbA1c levels between 5.7-6.4%, including 20 cases with previous DM diagnosis (Supplementary Table 2).

A total of 30/602 (5%) patients required hospital admission one month or later after the onset of disease. Most hospitalizations occurred between one to three months after disease onset, and were due to infection (n=14), pulmonary thromboembolism (n=3), dyspnea/chest pain (n=5), and tracheal stenosis (n=3) (Supplementary Table 3). Seven of the patients requiring later hospitalization were classified as having mild COVID-19 during the acute phase, while 23 were classified as moderate or severe.

The influence of PACS on patients’ quality of life was evaluated by EuroQoL (EQ-5D-5L) in 404/602 patients with different classifications of COVID-19 disease severity. Of these, 357 (88.4%) reported some degree of alteration, with 45 (11.1%) patients reporting inabilities or extreme values in at least one domain of the EQ-5D-5L. The most affected dimensions were anxiety/depression [79/401 (19.7%) reporting severe or extreme anxiety] and pain/discomfort [71/403 (17.6%) severe or extreme pain] (Supplementary Table 4). The median EuroQoL Global Score was 70 [IQR 50-80]. We further evaluated associations between clinical and demographic characteristics with respect to the impact on health-related quality of life. The female sex was associated with worse scores across all domains, except self-care. As expected, the domains of mobility and self-care were more affected in patients with severe acute disease. The most prevalent persistent symptoms (dyspnea, chest pain and fatigue) were found to be associated with decreased quality of life. In individuals reporting fatigue, all five domains of the EuroQol were negatively affected (Figure 3, Supplementary Table 5 and 6).

**Figure 3.**
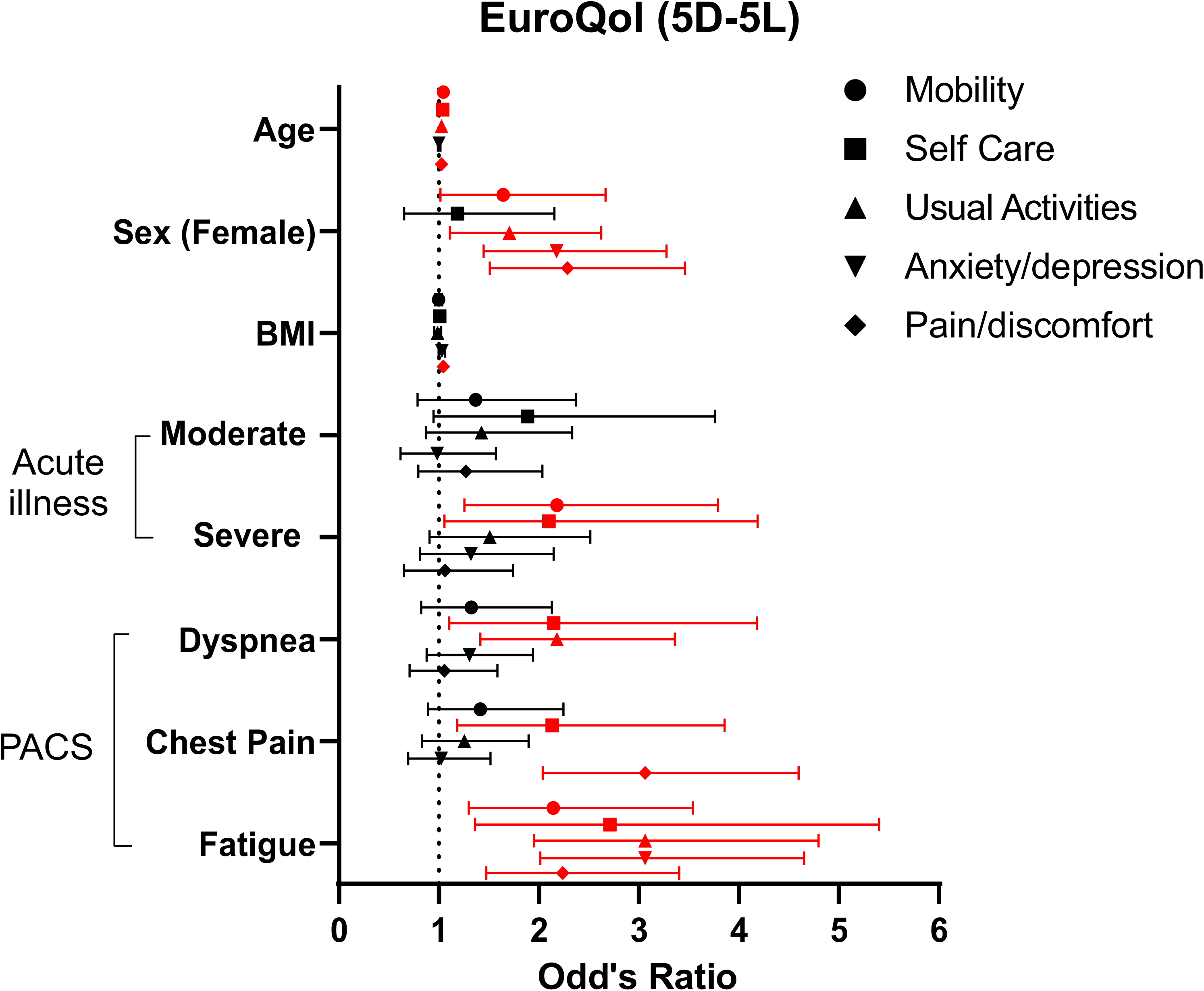
Forest plot showing the result of ordinal logistic regression for each domain of EuroQoL. Each symbol represents the odds ratio with the corresponding 95% confidence interval. Red line indicates *p* < 0.05.

## DISCUSSION

To the best of our knowledge, the present report represents an initial attempt to describe PACS in a sample of Latin American patients largely characterized as mixed-race or black. In addition to reinforcing findings reported by previous studies conducted in hospitalized patients from North America, Asia and Europe [15–18], similar persistent symptoms were noted among patients with PACS who were classified as having mild illness. Although PACS in patients with mild COVID-19 seemed to have less impact on quality of life, a small percentage (3%) did require hospitalization one month after disease onset, demonstrating that PACS after mild disease may also require active follow-up.

Regardless of disease severity, women presented more residual symptoms as evidenced by higher frequencies of fatigue, chest pain and myalgia, which stands in agreement with previous studies [3,15,19,20]. Women also reported worse quality of life scores in the domains of mobility, usual activities, anxiety/depression and pain. Long-term symptoms, as well as higher rates of anxiety and depression, have also been reported in females after infection with other viruses, including SARS-CoV. [21;22]. A recent report found that anxiety and depression were also associated with persistent fatigue in females six months after COVID-19 disease onset [3]. These conditions may exert bidirectional effects, since psychological disorders may exacerbate residual symptoms.

Dyspnea, fatigue, and chest pain were the most frequently reported symptoms. Reports of dyspnea are frequent in PACS patients, mainly after acute respiratory distress syndrome [4,23]. Our data indicate residual breathlessness regardless of disease severity during the acute stage and normal resting oxygen saturation. While acute disease have been linked to silent hypoxia (reduced pulse oximetry without breathless) [24,25], dyspnea may occur without detectable changes in pulse oximetry in patients with PACS. Apparently, post-acute dyspnea has a multifactorial etiology. In addition to lung disorders with abnormalities in chest computed tomography and lung function, anxiety, hyperventilation syndrome, respiratory muscle weakness and dysfunctional breathing have also been implicated [16,23,26–29]. The perception of breathlessness may be confused with or potentialized by other complaints. In 67.5% of our patients with dyspnea, fatigue was also reported - a common finding in PACS reported by 53 to 73% of SARS-CoV-2-affected patients [3,4,30]. Fatigue has also been reported at later times after SARS-CoV infection (in 64% of studied patients after three months[31] and in 40% after four years[32]). Immunological dysregulation is implicated in both dyspnea and fatigue, although chronic infection and iatrogenic complications following severe disease (e.g., long periods of immobility and corticosteroid use) are also recognized risk factors for muscle wasting and dysfunction [33,34, 37–39]. Our analysis indicated that both dyspnea and fatigue were related to reductions in quality of life; however, chest pain and myalgia, reported in 38 to 72%, were associated with the lowest EuroQol Global scores. It has been proposed that pain symptoms may be related to neuroinflammation, muscle lesion or direct viral injury [40]. Longstanding pain may require hospitalization for treatment and could lead to serious conditions that negatively impact patient health.

We identified 30 patients who required hospitalization at least one month after the onset of acute disease, mainly those who had previously been hospitalized. A recent British study that evaluated 47,780 individuals with an average follow-up of 140 days reported a readmission rate of 29.4%[17]. Hospitalization after discharge in patients with moderate to severe infection may be related to higher frequencies of comorbidities in this population[18,41]. Leijte *et al*.[42] reported an 11.7% rate of readmission in 769 patients followed for an average of 80 days; the risk factors identified were the male sex, discharge to a long-term care facility and COPD. Among the 23 patients herein with COPD seen at CPC, five (22%) were hospitalized later than one month after disease onset, suggesting that this clinical condition may be a risk factor for further complications. Surprisingly, among the 30 individuals who required late hospitalization, seven were classified as having mild COVID-19 (four were obese and two had hypertension). These admissions were mainly related to respiratory infection, chest pain or dyspnea.

DM is a known risk factor for severe COVID-19[43]. Here we observed increased levels of HbA1c in 184/280 PACS cases, most of whom had no previous diagnosis of DM. New onset of DM after COVID-19 has been postulated. SATHISH *et al*.[44] reported that 14.4% of 3,711 patients hospitalized due to COVID-19 presented new onset of DM. Huang *et al*.[3] reported 58 patients newly diagnosed with DM after discharge, one case presenting a normal level of HbA1c during hospitalization. Possible causes may be increased insulin resistance, elevated ACE2 levels, inflammation and autoimmune conditions. Prolonged glucocorticoid use can also increase HbA1c levels[45]. Importantly, this therapy was administered in 357 out of 493 (72.4%) of our PACS patients. Considering that HbA1c represents the mean glucose level during the last three months, value can be falsely enhanced by altered lifetime of red blood cells (such as in anemia related to iron deficiency)[46]. Indeed, anemia was detected in 48/184 patients who presented elevated HbA1c in our sample. Additionally, HbA1c level at acute stage was not available and previous undiagnosed DM should be considered. Future studies may compare HbA1c levels during hospitalization and late after discharge in order to clarify this important question.

Our study has several limitations, although sleep disorders and neurological sequelae has been extensively reported in PACS, we did not address these sequelae. The CPC is located in a reference hospital for respiratory diseases, and cases with respiratory complaints may have been preferentially referred to this health unit, therefore cases of mild COVID-19 may not be representative of outpatients with PACS. Additionally, it is not possible to compare PACS on mild and moderate/severe cases due to differences in the recruitment strategy. Hospitalized patients were referred to the CPC after discharge while outpatients attended by spontaneous demand, probably selecting patients with more severe residual symptoms and/or with comorbidities. Indeed, 53% of patients with mild disease reported at least one comorbidity.

In conclusion, we described PACS among hospitalized and non-hospitalized patients from Brazil three months following SARS-CoV-2 infection with a few cases of complications requiring hospitalization late after disease onset. Similar lingering and debilitating symptoms were detected in the spectrum of disease severity. Morbidity due to painful and physical-respiratory symptoms negatively affects quality of life, mainly in women who seem to be more affected regardless of disease severity. Considering the impact of PACS in daily activities and the high incidence of COVID-19 in Brazil, further studies should be performed to evaluate the magnitude of the problem and its impact on the public health system.

## Data Availability

The data that support the findings of this study are available from the corresponding author, MCCL, upon reasonable request.

## CPC study group

Ana Paula Andrade Barreto, Lucimeire Cardoso Duarte, Thiago Cerqueira-Silva, Marcio Andrade Barreto Filho, Aquiles Camelier, Natalia Tavares Machado, Manoel Barral-Netto, Viviane Sampaio Boaventura, Marcelo Chalhoub Coelho Lima, Maria de Fátima Fernandes Estrela, Mariângela Carneiro Ramos, Cristiane Leite Mesquita, Marizete Antunes Matos, Manoela Souza Trindade Fontes, Juliane Penalva Costa Serra, Maristela Sestelo, Marcia Maria Magalhães de Almeida e Marinho, Patrícia Alves Portela Santos, Carolina Neves, Eliana Dias Matos, Fabiane Costa Santos Fontoura, Thaiana Marcelino Ramos, Yasmin Pimenta Cadidé, Anna Lúcia Diniz, Kristine Menezes Barberino Mendes, Juliana Lemos de Sant’ana, Andréa Ferreira de Jesus, Marta França Santos, Carla Fernanda Rodrigues Guedes, Joelma Pereira da Conceição Anunciação, Vânia Lúcia de Sales Pedreira, Margaranei Vasconcelos Reis, Monica Suzart Gomes, Emília Augusta Franz Vieira Passos, Priscila Mary dos Santos Bahia, Milena Nogueira Azevedo, Giancarla Libório Di Credico

## Contributors

VSB, MBN, MCCL,AC conceived, designed, and supervised the study. TCS performed the formal analysis. APAB, TCS, MABF were responsible for data management. APAB, LCD, TCS, MBAF, VSB interpreted the data, and wrote the first draft. AC, NTM, MBN, MCCL critically revised the manuscript for important intellectual content. MCCL is the guarantor. The corresponding author attests that all listed authors meet authorship criteria and that no others meeting the criteria have been omitted.

## Acknowledgments

The authors are grateful to the involved in the patient’s clinical care, the nursing and administrative technicians Adriele Bastos, Eliene Gibaut, Alisson de Andrade, Antônio Luiz da Silva, Maria de Fátima da Silva, Denise Albuquerque, Estela Cordeiro, Jamile Oliveira, Monique Oliveira, Valcelia Muniz and Juliane Simões. We thank all patients who participated in this study and their families. The authors also thank Andris K. Walter for English language revision and manuscript copy editing assistance.

---

**Supplementary Table 1.**
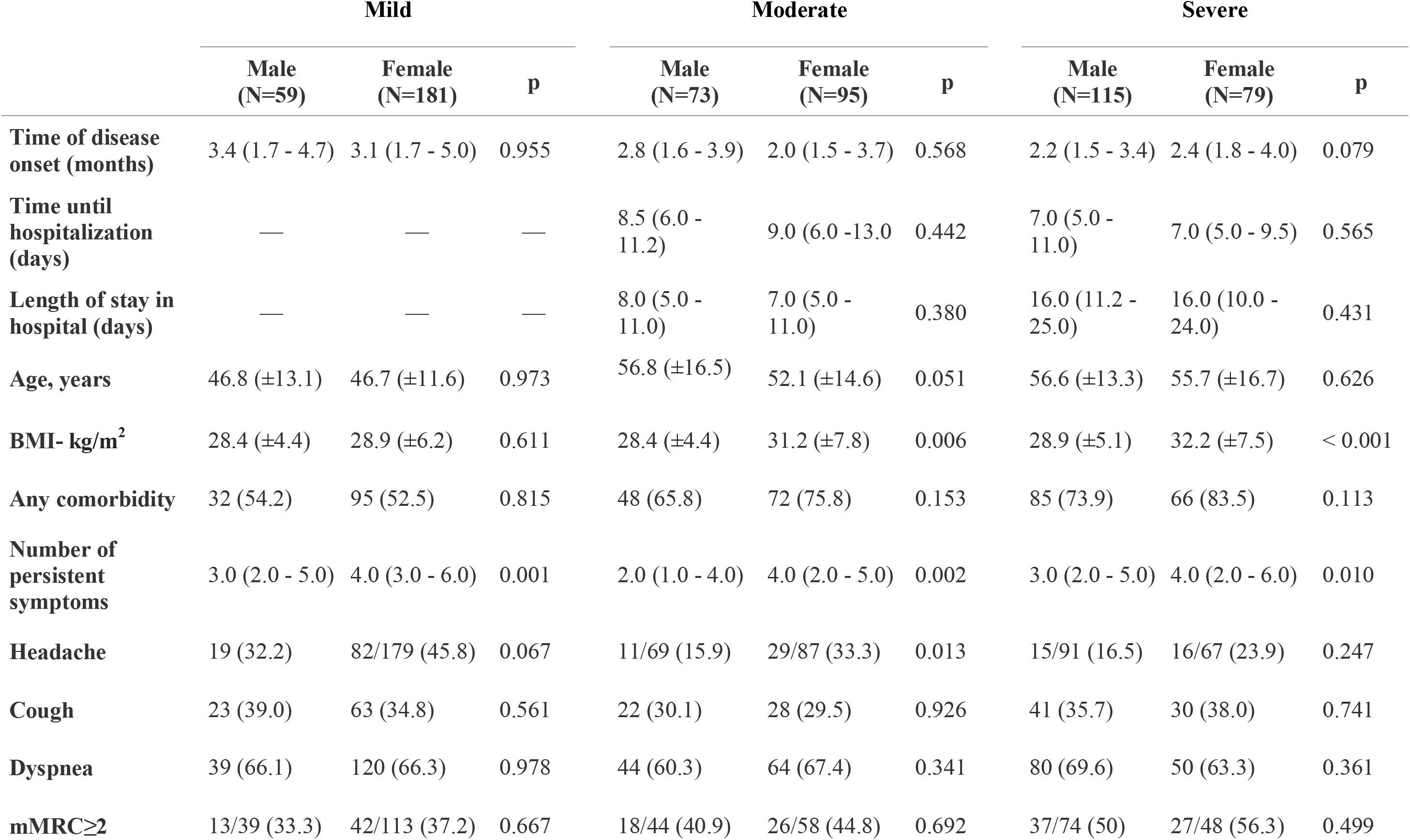
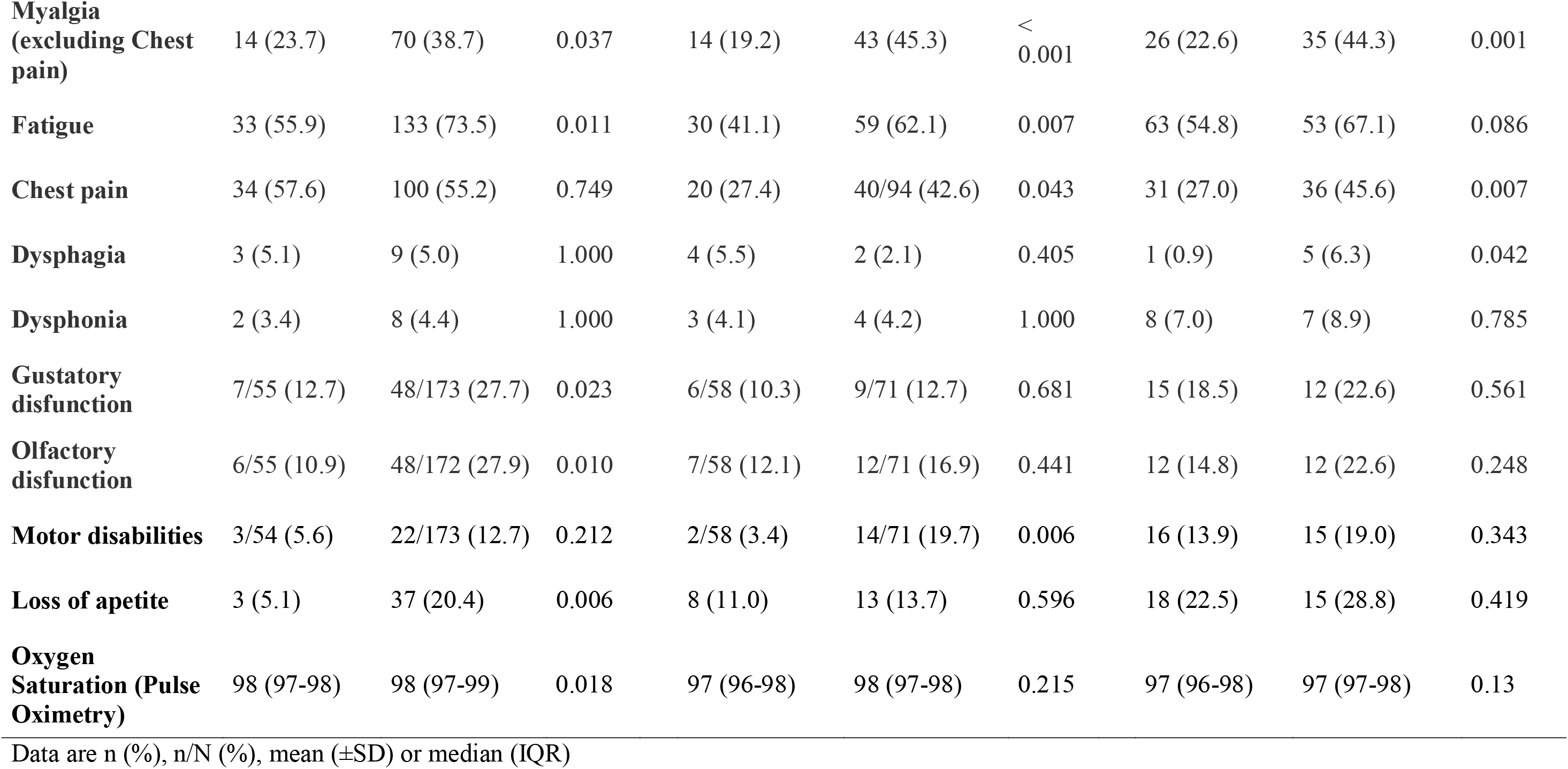
Persistent symptoms separated by sex and disease severity at the acute phase.

**Supplementary Table 2.** Laboratory findings of blood samples collected during admission

|  | n/N (%) |
| --- | --- |
| **Hemoglobin (<14/12 g/dL)\*** | 143/545 (26.2) |
| **Alanine Transaminase ( >72/52 U/L)\*** | 16/525 (3.0) |
| **Aspartate Transaminase (>59/36 U/L)\*** | 25/524 (4.8) |
| **Platelet (<14.000/dl or >450.000/dl)** | 30/544 (5.5) |
| **Urea (<19/15 mg/dL or >43/36 mg/dL)\*** | 88/536 (16.4) |
| **Creatinine (>0,8/0,7mg/dL or <1,5/1,2 mg/dL)\*** | 10/536 (1.9) |
| **White Blood Cell count per µl (> 3500 or < 10500)** | 65/542 (12.0) |
| **Lymphocytes count per µl ( >700 or <4410)** | 11/540 (2.0) |
| **Creatinine Phospokinase (>170/135 U/L)\*** | 67/497(13.5) |
| **Protein C Reactive ( >5mg/L)** | 195/484 (40.3) |
| **Potassium (<3,5 or >5,0 mEq/L)** | 59/512 (11.5) |
| **Sodium (<135 or >150 mEq/L)** | 19/513 (3.7) |
| **Glycated Hemoglobin\*\*** |  |
| <5.7% | 91 (33.1) |
| 5.7-6.4% | 108 (39.3) |
| >6.4% | 76 (27.6) |

\* Reference values for male and female, respectively
\*\* N=275

**Supplementary Table 3.** Characteristics of patients hospitalized one month or later after disease onset.

|  | **Outpatient**<br>**(N=7)~3%** | **Hospitalized**<br>**(N=23)~6%** |
| --- | --- | --- |
| **Age, years** | 42.5 (±4.9) | 56.6 (±18.4) |
| **Female** | 5 (71.4) | 8 (34.8) |
| **ICU admission acute phase** | — | 13 (56.5) |
| **Time after disease onset** |  |  |
| **1-3 months** | 5 (71.4) | 16 (69.6) |
| **>3 months** | 2 (28.6) | 7 (30.4) |
| **Obesity (BMI≥30 kg/m<sup>2</sup>)** | 4 (57.1) | 5 (21.7) |
| **Systemic Hypertension** | 2 (28.6) | 12 (52.2) |
| **Diabetes Mellitus** | 0 (0.0) | 7 (30.4) |
| **COPD** | 0 (0.0) | 5/22 (22.8) |
| **Cardiopathy** | 0 (0.0) | 4 (17.4) |
| **Bronchial asthma** | 0 (0.0) | 4 (17.4) |
| **Neoplasia** | 0 (0.0) | 3 (13.1) |
| **Chronic kidney disease** | 0 (0.0) | 1 (4.3) |
| **Cigarette smoking (former)** | 0 (0.0) | 8 (34.8) |
| **Cause of Hospitalization** |  |  |
| **Infection** | 2 (28.6) | 12 (52.2) |
| **Pulmonary thromboembolism** | 0 (0.0) | 3 (13.1) |
| **Tracheal stenosis** | 0 (0.0) | 3 (13.1) |
| **Chest pain** | 1 (14.3) | 2 (8.7) |
| **Dyspnea** | 1 (14.3) | 1 (4.3) |
| **Others** | 3 (42.9) | 2 (8.6) |

Data are n (%) or mean (±SD)

**Supplementary Table 4.** Descriptive EuroQol results separated by sex and disease severity at the acute phase.

|  | Mild |  |  | Moderate |  |  | Severe |  |  |
| --- | --- | --- | --- | --- | --- | --- | --- | --- | --- |
|  | Male<br>(N=48) | Female<br>(N=154) | p value | Male<br>(N=43) | Female<br>(N=56) | p value | Male<br>(N=61) | Female<br>(N=42) | p value |
| **EuroQoL Global Score\*** | 65.2<br>(±19.6) | 60.8 (±18.7) | 0.165 | 71.0 (±21.9) | 65.2 (±21.0) | 0.197 | 66.8 (±22.3) | 68.2 (±16.0) | 0.734 |
| **Anxiety** | N=47 | N=153 | 0.005 | N=43 | N=55 | 0.06 | N=61 | N=42 | 0.018 |
| No anxiety/depression | 20 (42.6) | 32 (20.9) |  | 22 (51.2) | 14 (25.5) |  | 23 (37.7) | 8 (19.0) |  |
| Slight anxiety/depression | 13 (27.7) | 30 (19.6) |  | 6 (14.0) | 12 (21.8) |  | 8 (13.1) | 12 (28.6) |  |
| Moderate anxiety/depression | 10 (21.3) | 55 (35.9) |  | 11 (25.6) | 15 (27.3) |  | 22 (36.1) | 9 (21.4) |  |
| Severe anxiety/depression | 4 (8.5) | 26 (17.0) |  | 4 (9.3) | 12 (21.8) |  | 4 (6.6) | 8 (19.0) |  |
| Extreme anxiety/depression | 0 (0) | 10 (6.5) |  | 0 (0.0) | 2 (3.6) |  | 4 (6.6) | 5 (11.9) |  |
| **Mobility** | N=48 | N=154 | 0.22 | N=43 | N=56 | 0.047 | N=61 | N=42 | 0.714 |
| No problems with walking around | 39 (81.2) | 105 (68.2) |  | 33 (76.7) | 33 (58.9) |  | 32 (52.5) | 20 (47.6) |  |
| Slight problems with walking around | 7 (14.6) | 23 (14.9) |  | 3 (7.0) | 11 (19.6) |  | 13 (21.3) | 9 (21.4) |  |
| Moderate problems with walking around | 1 (2.1) | 20 (13.0) |  | 4 (9.3) | 5 (8.9) |  | 10 (16.4) | 9 (21.4) |  |
| Severe problems with walking around | 1 (2.1) | 4 (2.6) |  | 1 (2.3) | 7 (12.5) |  | 2 (3.3) | 3 (7.1) |  |
| Unable to walk around | 0 (0.0) | 2 (1.3) |  | 2 (4.7) | 0 (0.0) |  | 4 (6.6) | 1 (2.4) |  |
| **Pain/Discomfort** | N=48 | N=154 | 0.008 | N=43 | N=56 | 0.003 | N=60 | N=42 | <0.001 |
| No pain/discomfort | 23 (47.9) | 40 (26.0) |  | 21 (48.8) | 10 (17.9) |  | 30 (50.0) | 9 (21.4) |  |
| Slight pain/discomfort | 12 (25.0) | 28 (18.2) |  | 9 (20.9) | 18 (32.1) |  | 11 (18.3) | 8 (19.0) |  |
| Moderate pain/discomfort | 11 (22.9) | 57 (37.0) |  | 10 (23.3) | 13 (23.2) |  | 5 (8.3) | 17 (40.5) |  |
| Severe pain/discomfort | 2 (4.2) | 24 (15.6) |  | 3 (7.0) | 15 (26.8) |  | 8 (13.3) | 7 (16.7) |  |
| Extreme pain/discomfort | 0 (0.0) | 5 (3.2) |  | 0 (0.0) | 0 (0.0) |  | 6 (10.0) | 1 (2.4) |  |
| **Self Care** | N=48 | N=154 | 0.353 | N=42 | N=56 | 0.298 | N=61 | N=42 | 0.854 |
| No problems with washing or dressing | 45 (93.8) | 128 (83.1) |  | 36 (85.7) | 42 (75.0) |  | 43 (70.5) | 33 (78.6) |  |
| Slight problems with washing or dressing | 2 (4.2) | 14 (9.1) |  | 3 (7.1) | 6 (10.7) |  | 7 (11.5) | 4 (9.5) |  |
| Moderate problems with washing or dressing | 0 (0.0) | 7 (4.5) |  | 1 (2.4) | 3 (5.4) |  | 6 (9.8) | 3 (7.1) |  |
| Severe problems with washing or dressing | 1 (2.1) | 3 (1.9) |  | 0 (0.0) | 4 (7.1) |  | 1 (1.6) | 0 (0.0) |  |
| Unable to wash or dress | 0 (0.0) | 2 (1.3) |  | 2 (4.8) | 1 (1.8) |  | 4 (6.6) | 2 (4.8) |  |
| **Usual Activities** | N=48 | N=153 | 0.093 | N=42 | N=56 | <0.001 | N=61 | N=42 | 0.883 |
| No problems with usual activities | 29 (60.4) | 67 (43.8) |  | 29 (67.4) | 17 (30.9) |  | 29 (47.5) | 18 (42.9) |  |
| Slight problems with usual activities | 7 (14.6) | 39 (25.5) |  | 5 (11.6) | 14 (25.5) |  | 13 (21.3) | 7 (16.7) |  |
| Moderate problems with usual activities | 9 (18.8) | 36 (23.5) |  | 2 (4.7) | 15 (27.3) |  | 9 (14.8) | 8 (19.0) |  |
| Severe problems with usual activities | 3 (6.2) | 4 (2.6) |  | 4 (9.3) | 8 (14.5) |  | 3 (4.9) | 2 (4.8) |  |
| Unable to do usual activities | 0 (0.0) | 7 (4.6) |  | 3 (7.0) | 1 (1.8) |  | 7 (11.5) | 7 (16.7) |  |

Data are n (%) or mean ( ± SD)
\*Only
patients

**Supplementary Table 5.** Ordinal Logistic regression for EuroQoL domains

|  | Mobility |  |  | Self-Care |  |  | Usual Activities |  |  | Anxiety/depression |  |  | Pain/discomfort |  |  |
| --- | --- | --- | --- | --- | --- | --- | --- | --- | --- | --- | --- | --- | --- | --- | --- |
|  | Odds Ratio | 95% CI | p value | Odd's Ratio | 95% CI | p value | Odd's Ratio | 95% CI | p value | Odd's Ratio | 95% CI | p value | Odd's Ratio | 95% CI | p value |
| **Fatigue** | 2.14 | 1.30 - 3.54 | 0.003 | 2.71 | 1.36 - 5.4 | 0.005 | 3.06 | 1.95 - 4.8 | <0.001 | 3.06 | 2.01 - 4.65 | <0.001 | 2.24 | 1.47 - 3.4 | <0.001 |
| **Chest pain** | 1.41 | 0.89 - 2.25 | 0.142 | 2.13 | 1.18 - 3.86 | 0.012 | 1.25 | 0.83 - 1.89 | 0.282 | 1.02 | 0.69 - 1.52 | 0.911 | 3.06 | 2.04 - 4.6 | <0.001 |
| **Dyspnea** | 1.32 | 0.82 - 2.13 | 0.251 | 2.15 | 1.10 - 4.18 | 0.025 | 2.18 | 1.41 - 3.36 | 0.004 | 1.3 | 0.88 - 1.94 | 0.192 | 1.06 | 0.70 - 1.58 | 0.792 |
| **Severe Acute illness** | 2.18 | 1.25 - 3.79 | 0.006 | 2.1 | 1.05 - 4.19 | 0.035 | 1.51 | 0.90 - 2.51 | 0.115 | 1.32 | 0.81 - 2.15 | 0.266 | 1.06 | 0.65 - 1.74 | 0.814 |
| **Moderate Acute illness** | 1.37 | 0.79 - 2.37 | 0.269 | 1.89 | 0.95 - 3.76 | 0.072 | 1.43 | 0.87 - 2.33 | 0.159 | 0.98 | 0.61 - 1.57 | 0.937 | 1.27 | 0.79 - 2.03 | 0.319 |
| **BMI-kg/m<sup>2</sup>** | 1 | 0.96 - 1.03 | 0.814 | 1.01 | 0.96 - 1.05 | 0.813 | 0.99 | 0.95 - 1.02 | 0.441 | 1.03 | 1.00 - 1.06 | 0.084 | 1.04 | 1.01 - 1.08 | 0.013 |
| **Sex (Female)** | 1.64 | 1.01 - 2.67 | 0.044 | 1.18 | 0.65 - 2.16 | 0.582 | 1.7 | 1.11 - 2.62 | 0.015 | 2.18 | 1.45 - 3.28 | <0.001 | 2.29 | 1.51 - 3.46 | <0.001 |
| **Age, years** | 1.04 | 1.03 - 1.06 | <0.001 | 1.04 | 1.01 - 1.06 | <0.001 | 1.02 | 1.01 - 1.04 | 0.001 | 1 | 0.99 - 1.02 | 0.868 | 1.02 | 1.01 - 1.04 | 0.001 |

**Supplementary Table 6.** Multiple linear regression for EuroQoL Global Score

|  | **Global** |  |  |
| --- | --- | --- | --- |
|  | Beta (Estimate) | 95% CI | p value |
| **Fatigue** | -8.28 | -12.5 to - 4.05 | <0.001 |
| **Chest pain** | -8.62 | -12.76 to -4.49 | <0.001 |
| **Dyspnea** | -4.96 | -9.1 to -0.83 | 0.019 |
| **Severe** | 3.98 | -1.08 to 9.05 | 0.123 |
| **Moderate** | 2.67 | -2.21 to 7.55 | 0.283 |
| **BMI- kg/m<sup>2</sup>** | -0.02 | -0.34 to 0.34 | 0.99 |
| **Sex (Female)** | -0.73 | -4.94 to 3.48 | 0.732 |
| **Age, years** | -0.13 | -0.28 to 0.01 | 0.077 |

